# Municipality- level predictors of COVID-19 mortality in Mexico: a cautionary tale

**DOI:** 10.1101/2020.07.11.20151522

**Authors:** Alejandra Contreras-Manzano, Carlos M. Guerrero-López, Mercedes Aguerrebere, Ana Cristina Sedas, Héctor Lamadrid-Figueroa

**Affiliations:** Independent consultant, Mexico; Mexican Social Security Institute, Mexico; National Autonomous University of Mexico, Mexico; Department of Global Health and Social Medicine, Harvard Medical School, United States of America; Center for Population Health Research, National Institute of Public Health, Mexico

**Keywords:** COVID-19, risk factors, social determinants, health determinants, Municipality-level, counties

## Abstract

**Background:** Inequalities and burden of comorbidities of the Coronavirus disease 2019 (COVID-19) vary importantly inside the countries. We aimed to analyze the Municipality-level factors associated with a high COVID-19 mortality rate of in Mexico.

**Methods:** We retrieved information from 142,643 cumulative confirmed symptomatic cases and 18,886 deaths of COVID-19 as of June 20^th^, 2020 from the publicly available database of the Ministry of Health of Mexico. Public official data of the most recent census and surveys of the country were used to adjust a negative binomial regression model with the quintiles (Q) of the distribution of sociodemographic and health outcomes among 2,457 Municipality-level. Expected Mortality Rates (EMR), Incidence Rate Ratios (IRR) and 95% Confidence Intervals are reported.

**Results:** Factors associated with high MR of COVID-19, relative to Quintile 1 (Q1), were; diabetes prevalence (Q4, IRR=2.60), obesity prevalence (Q5, IRR=1.93), diabetes mortality rate (Q5, IRR=1.58), proportion of indigenous population (Q2, IRR=1.68), proportion of economically active population (Q5, IRR=1.50), density of economic units that operate essential activities (Q4, IRR=1.54) and population density (Q5, IRR=2.12). We identified 1,351 Municipality-level without confirmed COVID-19 deaths, of which, 202 had nevertheless high (Q4, Mean EMR= 8.0 deaths per 100,000) and 82 very high expected COVID-19 mortality (Q5, Mean EMR= 13.8 deaths per 100,000).

**Conclusion:** This study identified 1,351 Municipality-level of Mexico that, in spite of not having confirmed COVID-19 deaths yet, share characteristics that could eventually lead to a high mortality scenario later in the epidemic and warn against premature easing of mobility restrictions. Local information should be used to reinforce strategies of prevention and control of outbreaks in communities vulnerable to COVID-19.

**Key messages:**

- Predictors of COVID-19 mortality varied importantly between Municipality-level.
- Municipality-level factors associated with high mortality of COVID-19 were the prevalence of obesity and diabetes, mortality rate of diabetes, the proportion of indigenous and economically active population and population density.
- Municipality-level with high case-fatality rates of COVID-19 are likely undergoing insufficient testing and should improve its availability.
- Identified predictors ought to be considered by local governments to reinforce tailored strategies to prevent casualties in populations vulnerable to COVID-19, as mortality is expected to be eventually high in some Municipality-level that may not have reached the apex of the epidemic yet.

## Introduction

The first case of the new Coronavirus disease 2019 (COVID-19) in Mexico was confirmed on February 28^th^, 2020^1^. Since then, the Government of Mexico has launched a series of preventive measures which adhere to the World Health Organization (WHO) SARS-Cov-2 strategic preparedness and response plan aimed at limiting the spread of the virus^2^. The “National Campaign for Healthy Distance”, implemented from March 23^rd^ to May 30^th^ 2020 included social distance, hand washing, general confinement, self-isolation for those with COVID-19 associated symptoms for 14 days and limited economic mobility^3^. As of June 20^th^, the spread of the virus continued to rise, resulting in 175 148 accumulated positive cases and 20 773 deaths^4^.

Globally, individual factors associated with COVID-19 mortality have been found to be: male sex, age over 65 years, ethnicity, hypertension, diabetes, cardiovascular disease, and respiratory disease, among others^5,6^. According to the 2018 National Health and Nutrition Survey (ENSANUT-2018) of Mexico, 36.1% of adults over the age of 19 were obese, 39.1% overweight, 13.7% diabetic and 25% hypertensive^7^. Considering that most states in Mexico hold a heterogeneous distribution in the prevalence of such comorbidities, regions with an increased burden of these diseases and sociodemographic related factors are at higher risk of encountering more severe manifestations of COVID-19 which might require hospitalization or critical care, as well as higher COVID-19 mortality rates.

It is deemed that in order to curb the pandemic, national decisions ought to be in coordination with those at a local level. In the United States sociodemographic and health factors related to COVID-19 vary importantly across counties^8-10^. Mexico has 2,457 Municipalities distributed in 32 states, which are the basis of the territorial organization and the political and administrative division^11^. By June 1^st^, 2020, the federal government gave the states of the country the responsibility for deciding the reopening of social, educational and economic activities based on a “Traffic Light” system established by the MoH^12^. COVID-19 is however expected to linger within communities for several months or even years to come. Targeted public policy interventions in regions that are highly vulnerable to COVID-19 are crucial to safeguard, protect and strengthen communities facing the pandemic. Therefore, the objective of this study was to analyze the Municipality-level factors associated with a higher mortality rate (MR) of COVID-19 in Mexico, and to pinpoint locations expected to suffer from a high COVID-19 mortality.

## Methods

Coronavirus pandemic surveillance in Mexico has been carried out using the Sentinel model proposed by the Pan American Health Organization (PAHO)^13^. This model works with 475 nationally representative health facilities that monitor coronavirus cases through the test for SARS-CoV-2 conducted in ten percent of suspected cases and 100% those suspected with severe acute respiratory syndrome and signs of breathing difficulty, or in deaths of those hospitalized suspected to be COVID-19 cases. The operational definition of a suspected case in Mexico is of an individual who in the last 7 days presented at least 2 of the following signs and symptoms: cough, fever, or headache, accompanied by at least one of the following signs or symptoms: dyspnea, arthralgia, myalgia, odynophagia/pharyngeal burning, rhinorrhea, conjunctivitis and/or chest pain^14^. A confirmed case of COVID-19 is defined as a person with a diagnosis given by the National Network of Public Health Laboratories recognized by the Institute of Epidemiological Diagnosis and Reference (InDRE) who met the criteria of a suspected case^15^.

We used the daily updated open data source from the General Directorate of Epidemiology of Mexican MoH which includes demographic and health information of confirmed, negative, and suspect cases of COVID-19.^4^ We extracted the data as of June 20^th^, 2020 of the number of confirmed symptomatic cases and deaths.

At the Municipality-level level, we obtained data on variables that we hypothesized could be linked to differential testing practices or exposure to SARS-Cov-2 of the most recent and complete information from census, and records from the National Institute of Statistics and Geography (*INEGI* per the Spanish acronym) of the 2457 Municipality-level according to the 2017 geographical division of the country^16^. We selected the following variables and years^17-19^; population (2010 and 2015), illiteracy in population aged 15 years or more (2015), proportion of indigenous population of 5 years or more (2010), proportion of the population over 12 years of age who are economically active (2015), population with private health insurance(2015), population affiliated to the Mexican Social Security Institute (IMSS, per the Spanish acronym) (2015), population without health care insurance (2019), proportion of households with indoor availability of water service (2015), proportion of households without sewage drainage (2015), population density (2015), and rate of economic units that operate essential activities during COVID-19 outbreak in Mexico from the National Directory of Economic Units/INEGI^20^.

From the Department of Epidemiological Surveillance of the Ministry of Health (DGIS for its acronym in Spanish), we retrieved records of deaths due to diabetes, hypertension and cardiovascular disease in adults over 20 years of age in 2018^21^.

From the catalog of the Unique Key of Health Establishments (CLUES per the Spanish acronym) we retrieved the number of medical units per Municipality-level for 2020 and we calculated the rate per 100 thousand inhabitants in the Municipality for each type of medical unit^22^. Rates and proportions of the variables at Municipality-level level were calculated using the annual population projections from the National Population Council (CONAPO)^23^. We obtained the State level estimations for the prevalence of obesity, previously diagnosed diabetes and previously diagnosed hypertension among adults aged 20 years and over from the latest National Survey of Nutrition and Health (ENSANUT-2018) 24.

### Statistical analysis

To identify the Municipality-level factors associated with the mortality rate of COVID-19, we adjusted a negative binomial regression model in which the dependent variable was the sum of confirmed COVID-19 deaths in symptomatic cases (n=18 886) by Municipality of residence and the independent variables were the quintiles of the distribution of sociodemographic and health outcomes within the Municipality. Expected Mortality Rates (EMR) for each Municipality, Incidence Rate Ratios (IRR) and 95% Confidence Intervals were estimated.

To better understand the association between MR of COVID-19 and Municipality-level factors, and in addition to the main objective of the study, we also fitted negative binomial regression models with the cumulative incidence rate of confirmed symptomatic cases (n=142 643) [(cases/estimated population)*100 000 population], and with the case-fatality rate in confirmed deaths of COVID-19 (n=18 886 deaths) [(deaths/confirmed cases)*100] as of June 20, 2020. In a supplementary table results are displayed stratified by months since the beginning of the pandemic in Mexico.

Finally, to contrast our findings on mortality risk factors with those at individual level in Mexico, we retrieved the following variables from the MoH coronavirus database; age, sex, state, Municipality of residence, indigenous languages speaker, diabetes, obesity, hypertension, COPD, cardiovascular disease, chronic kidney disease (CKD), immunosuppression, asthma, the dates of symptom onset, the hospital admission, and death. We excluded 429 cases as they had incomplete information on the covariates above mentioned. We fitted a Poisson regression model where the dependent variable was the binary outcome of death due to COVID-19 and the independent variables were the characteristics and binary comorbidities of the individual. The analysis and maps were developed using the statistical program STATA 14 (Stata Statistical Software: Release 14. College Station, TX: StataCorp LP), findings at p<0.05 were considered significant.

## Results

From 402 083 observations, 35% were positive symptomatic cases of COVID-19, 50.3% negative and 14.2% suspect cases. The proportion of deaths was 11.8% in positive cases, 2.56% in negative cases, and 2.71% in suspect cases. At the moment of the analysis, two thirds of the Municipality-level had symptomatic cases and almost half had reported deaths due to COVID-19. (data not shown)

In Table 1 are displayed the means and 95% CI of the Municipal and State-level factors included in the study, source of information and year of reference.

**Table 1.** Means of the Municipal and State-level factors included in the study and sources of information.

|  |  |  |  |  |
| --- | --- | --- | --- | --- |
| Variable | Mean (95% CI) | Source of information | Year | Level |
| Proportion of population aged 60 year or older | 0.14 (0.14, 0.14) | CONAPO/INEGI | 2020 | Municipal |
| Proportion of males | 0.48 (0.48, 0.48) | CONAPO/INEGI | 2020 | Municipal |
| Prevalence of diabetes in adults aged 20 years or older | 0.1 (0.1, 0.1) | ENSANUT/INEGI | 2018 | State |
| Prevalence of obesity in adults aged 20 years or older | 0.35 (0.34, 0.35) | ENSANUT/INEGI | 2018 | State |
| Prevalence of hypertension in adults aged 20 years or older | 0.18 (0.18, 0.18) | ENSANUT/INEGI | 2018 | State |
| Crude mortality rate of diabetes in adults 20 years or older | 133.2 (129.5, 136.8) | DGIS/INEGI | 2018 | Municipal |
| Crude mortality rate of hypertension in adults 20 years or older | 15.7 (14.3, 17.2) | DGIS/INEGI | 2018 | Municipal |
| Crude mortality rate of cerebrovascular disease in adults 20 years or older | 22.3 (20.6, 24.1) | DGIS/INEGI | 2018 | Municipal |
| Proportion of the population in extreme poverty | 0.2 (0.19, 0.21) | CONEVAL | 2015 | Municipal |
| Proportion of illiterate population aged 5 year or older | 0.12 (0.11, 0.12) | INEGI | 2015 | Municipal |
| Proportion of indigenous population aged 5 year or older | 0.19 (0.18, 0.21) | INEGI | 2010 | Municipal |
| Proportion of population economically active aged 12 year or older | 0.41 (0.41, 0.42) | INEGI | 2015 | Municipal |
| Rate of economic units classified as essential during COVID-19 outbreak | 0.03 (0.03, 0.03) | INEGI | 2020 | Municipal |
| Population density | 292.7 (245.6, 339.7) | INEGI | 2015 | Municipal |
| Proportion of houses with water service inside the domicile | 0.07 (0.07, 0.07) | INEGI | 2015 | Municipal |
| Proportion of houses without sewage drainage service | 0.19 (0.18, 0.2) | INEGI | 2015 | Municipal |
| Proportion of houses with dirt floor | 0.08 (0.08, 0.09) | INEGI | 2015 | Municipal |
| Rate of Hospitals | 2.39 (2.21, 2.57) | CLUES catalog | 2020 | Municipal |
| Rate of units of Health Care Services | 58.8 (56.6, 61.0) | CLUES catalog | 2020 | Municipal |
| Rate of Social Assistance medical units | 3.65 (2.8, 4.49) | CLUES catalog | 2020 | Municipal |
| Proportion of population without health care insurance (was affiliated to the People's Health Care) | 0.64 (0.63, 0.64) | INEGI | 2019 | Municipal |
| Proportion of population affiliated to the Mexican Social Security Institute (IMSS) | 0.14 (0.14, 0.15) | INEGI | 2015 | Municipal |
| Proportion of population with Private Health Insurance | 0.01 (0.01, 0.01) | INEGI | 2015 | Municipal |

Municipality-level factors associated with high mortality of COVID-19 were the prevalence of diabetes (Quintile 4, IRR= 3.43, 95% CI; 1.75-2.98), and obesity (Quintile 5, IRR= 1.72, 95% CI; 1.20-2.47), the mortality rate of diabetes (Quintile 5, IRR= 1.49, 95% CI: 1.15-1.93), proportion of indigenous population (Quintile 4, IRR= 1.51, 95% CI; 1.20-1.91), proportion of economically active population (Quintile 5, IRR= 1.59, 95% CI; 1.09-1.32), and population density (Quintile 5, IRR; 2.5, 95% CI; 1.78-3.51). Factors inversely associated with lower mortality of COVID-19 at the Municipality-level were the hypertension prevalence (Quintile 5, IRR= 0.39, 95%CI; 0.29-0.52) and characteristics of marginalized populations such as; illiteracy (Quintile 5, IRR= 0.62, 95% CI; 0.40-0.95), houses without drainage (Quintile 5 IRR; 0.71, 95% CI; 0.51-0.90), houses with dirt floors (Quintile 5, IRR= 0.68, 95%CI; 0.47-1.00), proportion of population without health care insurance (Quintile 5, IRR= 0.66, 95%CI; 0.45-0.96). (Table 2)

**Table 2.**
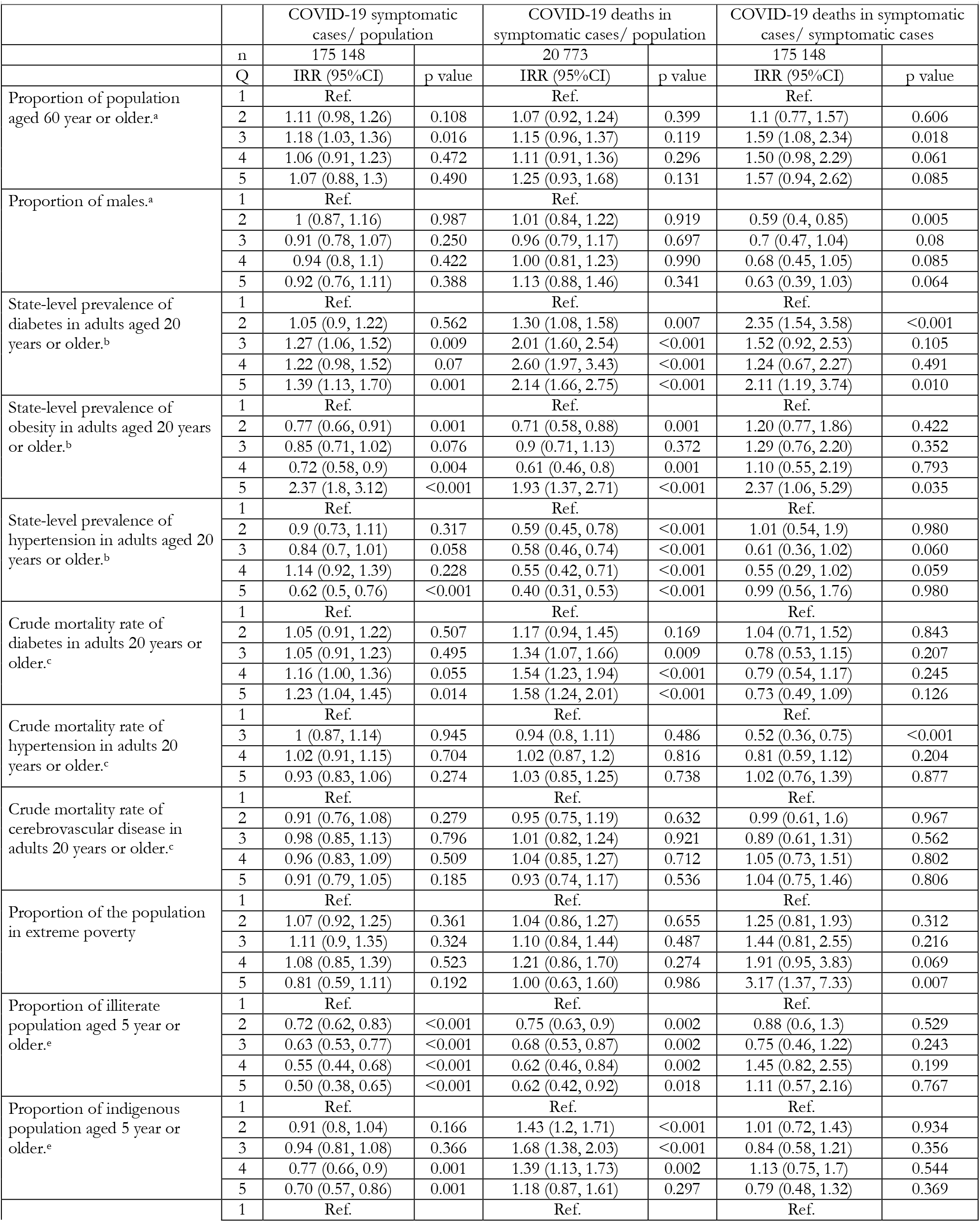
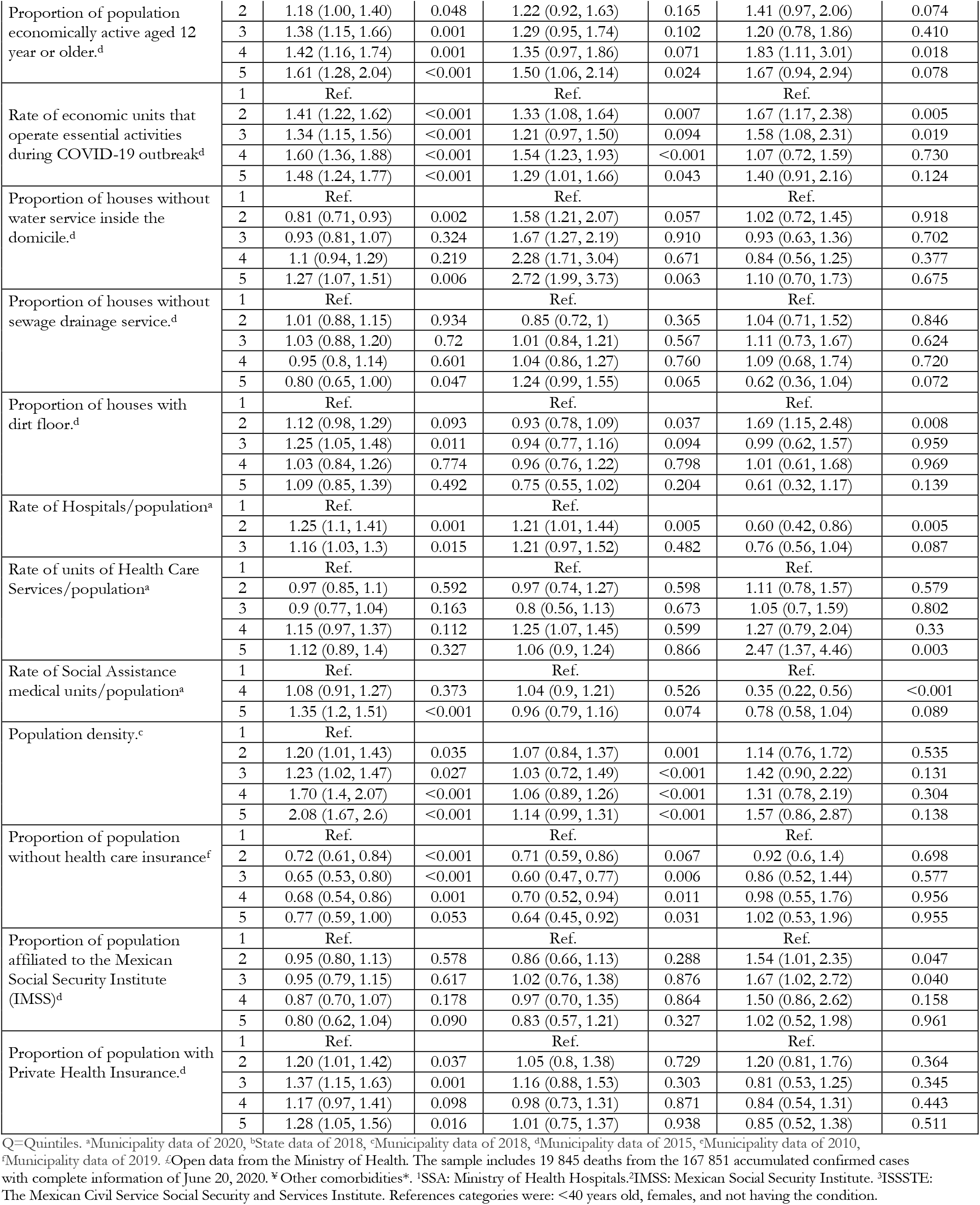
Municipal and State-level factors associated with higher cumulative incidence, mortality and case-fatality of COVID-19 in symptomatic cases in Mexico as of June 20, 2020.

Municipality-level factors associated with high cumulative incidence rate of COVID-19 were similar than those associated to high mortality, except by the proportion of indigenous population (Quintile 5, IRR; 0.70, 95%CI; 0.57-0.86) and the proportion of houses without sewage drainage (Quintile 5, IRR= 0.80; 0.65-1.00) which were inversely associated (Table 2). Finally, Municipality-level factors associated with higher case-fatality of COVID-19 were as well the prevalence of diabetes (Quintile 5, IRR= 2.11, 95%CI; 1.19-3.74), obesity prevalence (Quintile 5, IRR; 2.37, 95%CI; 1.06-5.29), extreme poverty (Quintile 5, IRR= 3.17, 95%CI; 1.37-7.33), and rate of social assistance medical units (Quintile 5, IRR; 2.47, 95%CI; 1.37-4.46). Quintiles of Crude mortality and expected mortality rates of COVID-19 at Municipality-level-level are shown in Map 1 and Map 2, respectively.

**Map 1.**
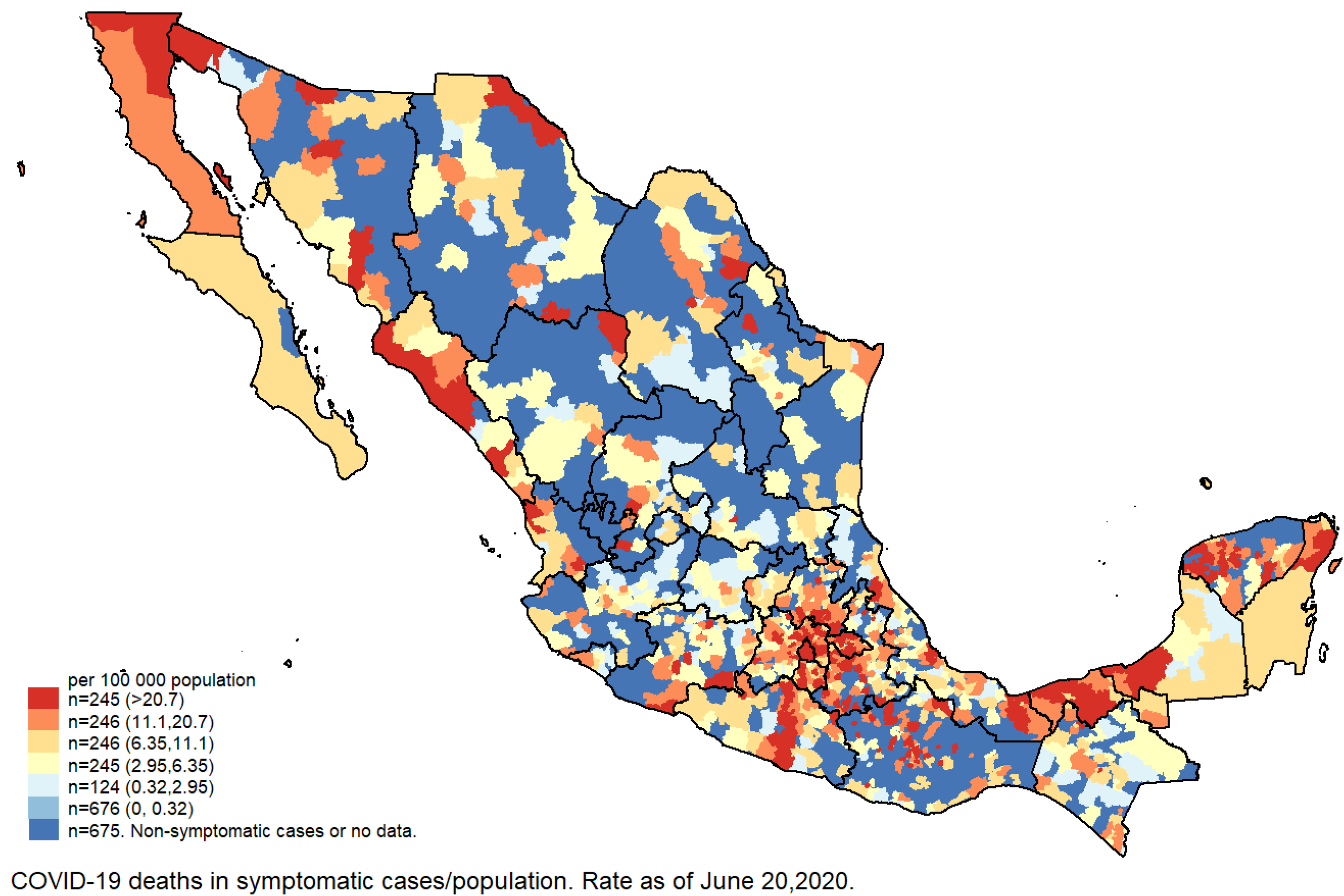
Crude Mortality Rate of symptomatic cases of COVID-19 in Mexico as of June 20, 2020.

**Map 2.**
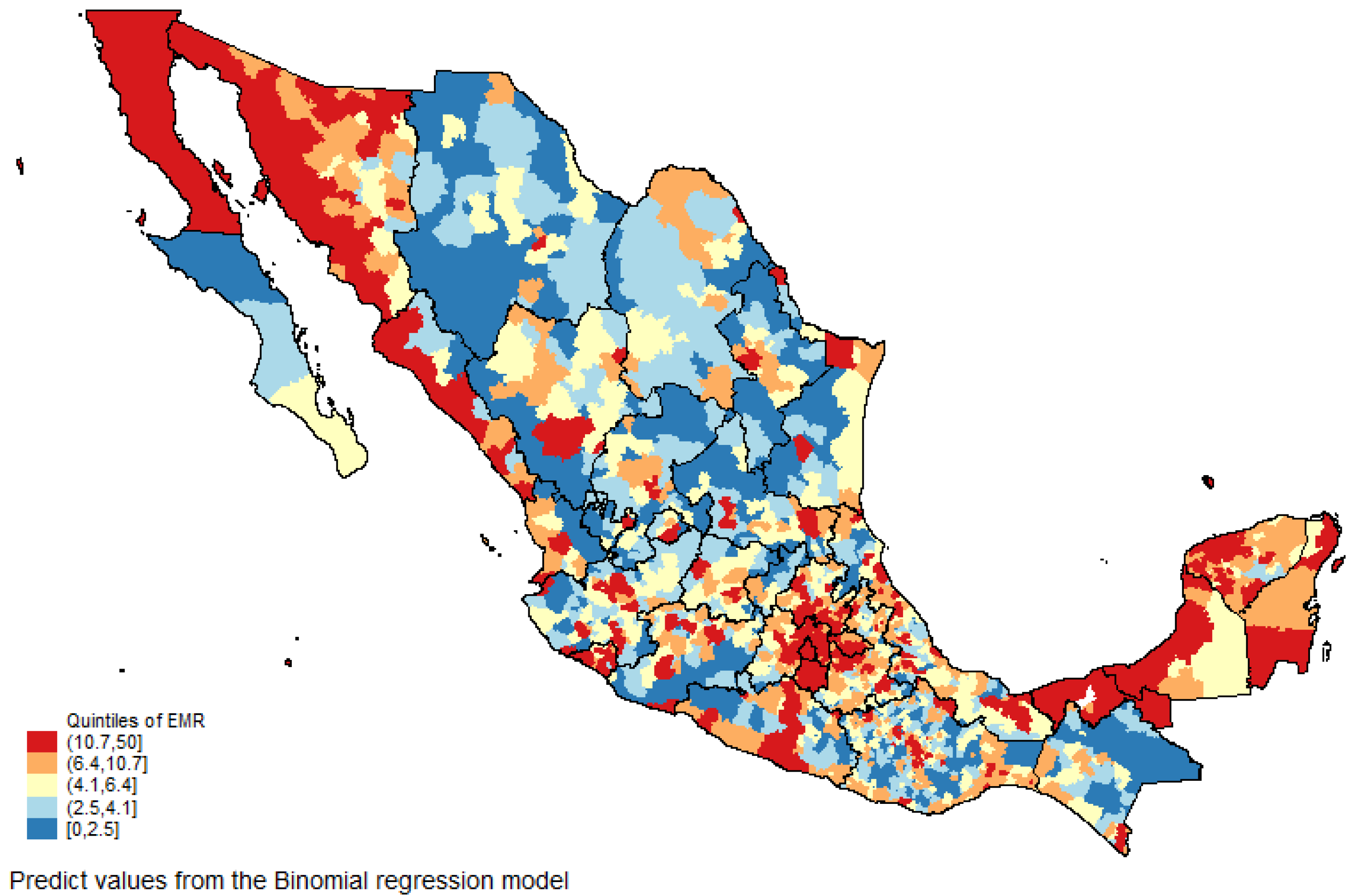
Expected Mortality Rate (EMR) of COVID-19 according to municipal factors studied.

We identified 1 351 Municipality-level without COVID-19 deaths reported in symptomatic cases, which according to its characteristics, 202 had high and 82 very high EMR of COVID-19 (Q4, Mean= 8.0, 95%CI: 5.7-11.2 and Q5, Mean= 13.8, 95%CI: 9.97-19.1 respectively). Supplementary Table 1 lists the name and characteristics of the Municipality-level with high expected mortality rate COVID-19 based on the significant risk factors identified in the binomial regression model (Table 2). It is observed that many Municipality-level of the states of Oaxaca (n=68), Yucatán (n=60), Sonora (n=33) Veracruz (n=25), Puebla (n=24) and Michoacán (n=18) had the highest expected mortality rates (Quintile 4, Mean= 8.01, 95%CI; 7.85-8.17) (Quintile 5, Mean= 13.7, 95%CI; 13.1-14.4). It is observed that 43 Municipality-level with high expected mortality of COVID-19 are Hope Municipality-level. Case-fatality rate of COVID-19 in symptomatic cases are shown in Supplementary Map 1. Can be observed that some Municipality-level of the States of Oaxaca, Chihuahua and Guerrero had case-fatality rates of COVID-19 higher than 80%.

At the individual level, we also explored characteristics of the confirmed symptomatic cases that were associated with higher lethality of COVID-19 in order to observed if the risk factors observed in other countries remain in the Mexican context. (supplementary Table 2.) In concordance with the Municipality-level results, diabetes, belonging to an indigenous ethnic group, and obesity were associated with higher risk of mortality due to coronavirus. Other individual risk factors of lethality-case were age 40 or over, male sex, hypertension, CKD, COPD and immunosuppression. In addition, having received medical care in private health institutions was associated with a lower lethality of COVID-19 compared to those cared for at public health services. As for individuals who sought medical attention between 6 and 14 days after the onset of symptoms, they had a higher risk of dying compared to those who received medical care between 0 and 5 days after the onset of symptoms.

In supplementary Table 3, we illustrate the Municipality-level factors associated to the cumulative incidence and lethality rates as of June 20^th^, 2020 and disaggregated by month. We found that Municipality-level factors association with cumulative incidence and mortality rate of COVID-19 varied within the course of the pandemic in Mexico.

## Discussion

Municipality-level factors found to be associated with higher risk of mortality of COVID-19 were the population density, the prevalence of diabetes and obesity, the mortality rate of diabetes, the proportion of indigenous population, of economically active population and the rate of economic units that operate essential activities during the COVID-19 outbreak.

First, high proportion of population economically active may reflect lower confinement and increased mobility, while higher population density is may be related to less social distancing and high connectivity. Secondly, high mortality rate of diabetes means inadequate control of glucose levels - which has a negative impact on the immune and cardiovascular systems both crucial in the body’s response to COVID-19^25^ - and also, lower access and quality of medical care, or weaker local health care systems^26^.

Despite of at the individual level, the hypertension was associated with higher lethality of COVID-19. At the Municipality level, the mortality rate of hypertension was not associated with high mortality of COVID-19, and within States with the highest prevalence of hypertension they had lower mortality risk of COVID-19. This result could be explained by the fact that the prevalence used in our model came from adults already diagnosed with hypertension, which according to previous surveys represented approximately 60% of the total cases of hypertensive population and are those with access to health care services and antihypertensive medication^27^. Nevertheless, unknown and unobserved variables, prior to the demand of healthcare services might be reflecting an inverse association between hypertension and COVID-19 mortality.

It is important to note that the cumulative incidence of cases in indigenous populations was low; however, the risk of mortality from coronavirus was higher than in Municipalities with lower proportion of indigenous population. These findings could indicate a lower access to healthcare services and testing, combined with a dynamic where the epidemic migrates further into more marginalized areas with a higher proportion of indigenous population. Other countries have found that many mechanisms underlying the higher lethality of COVID-19 in ethnic minorities are related to marginalization conditions such as locality of residence, work conditions and health inequalities^28,29^.

Marginalized conditions such as non-sewage service or dirt floors were inversely associated with mortality of COVID-19. Communities with these household characteristics are mostly remote and with low connectivity, which could be protective from SARs-cov-2 exposure. Notwithstanding, marginalized and remote communities could also have lower access to both testing for SARs-Cov-2 and health care which could lead to underreporting of cases and deaths. Special screening strategies and tracing and isolation of contacts within these vulnerable communities are essential to favor the prevention of deaths from COVID-19^30^.

In May 2020, the Mexican MoH published a list of 324 *Municipios de la Esperanza* (“Hope Municipalities”) which in the last 28 days prior to May 16 did not have any confirmed cases of COVID-19 and were not adjacent to municipalities with confirmed cases. This list was a guide for State governors to restart non-essential activities on May 18, 2020^31^. However, according to our results, 16% of these Municipality-level are at high risk of having worse outcomes in the event of an outbreak and, as of June 20, 2020, 389 symptomatic cases and 29 deaths due to COVID-19 were confirmed in these municipalities.

The identification of municipalities with a high burden of risk factors of severe illness or death of COVID-19 is critical to establish the most convenient health policies at the Municipality level. Many of these municipalities are in locations susceptible to an outbreak onset as the pandemic evolves in Mexico. municipalities with no confirmed cases yet, but with populations at risk of becoming seriously ill from COVID-19 could have a greater burden of the disease in the upcoming months if contingency measures and mobility restrictions are eased to soon. A targeted approach will be crucial to prevent or control the onset of new outbreaks within these municipalities include testing, isolation and contact tracing as well as more general measures according to the three-stage traffic light system that was established in Mexico to reactivate the economy and reduce contingency measures^12^.

Additional to the Municipality-level factors associated with the cumulative incidence and mortality of COVID-19 we identified that Municipalities in condition of extreme poverty had higher case-fatality rate of COVID-19. For example, Oaxaca, Chihuahua and Guerrero are States in which some municipalities had case-fatality rates higher than 80%. This finding suggests a low availability of tests in this communities.

As per the limitations within our study, que precision of the estimations depends on the quality of the databases for example; the aging of our data for some variables is up to 10 years, which may not reflect heterogeneous changes within municipalities by 2020. Nevertheless, data from previous years, still holds relation to the following years and they were useful to find expected associations. For obesity, diabetes and hypertension prevalence, the information from the ENSANUT-2018 are representatively at State level reducing the variability within Municipalities. However, for the rest of the variables, such as mortality rates of diabetes and hypertension, information was disaggregated at the municipality. Finally, the ecological design of our study prevented us to from establishing causal associations.

To our knowledge this is the first study that estimated mortality rate of COVID-19 by using the burden of related comorbidities and sociodemographic characteristics, in order to identify municipalities at risk of high mortality rates of coronavirus in Mexico. Our findings could contribute to the national strategic preparedness and response plans towards a “new normality”^30^ by informing at a Municipality-level level factors ought to consider in the decision-making process and public health interventions to minimize the negative impact of COVID-19 on the health and livelihoods of the most at-risk communities.

Based on our results, we considered this is a good moment to modify the current epidemiological surveillance strategy for confirming COVID-19 positive cases and deaths in populations who might be under-reported by the Sentinel surveillance approach. For instance, indigenous communities and communities with extreme poverty could be affected not only by the risks of COVID-19 afflicting the health of the population, but also by increasing food insecurity, domestic violence, disrupting the routine care of chronic diseases or the economic repercussions this might bring. It is critical to count on data on the impact of COVID-19 amongst these populations to identify, prioritize and address the needs of these vulnerable populations.

## Conclusion

Using small area demographic characteristics and burden of comorbidities of COVID-19 is useful to identify locations at risk of COVID-19 mortality. In Mexico, Municipality-level risk factors associated with high mortality rates of COVID-19 were high proportion of the population economically active, high population density, high proportion of indigenous population, and high diabetes mortality. Based on their characteristics many of the municipalities that have not experienced high mortality yet are prone to do so as the epidemic curve progresses. We therefore warn against overconfidence and premature easing of mobility restrictions and other contingency measures. Local governments ought to reinforce local strategies to prevent outbreaks in vulnerable communities to COVID-19.

## Data Availability

Data bases used in the study were retrieved from the publicly available databases of the Ministry of Health and public official data of the most recent census and surveys of Mexico.

https://www.dropbox.com/sh/cyrx2ly5vxf3a13/AAC6vBtZTe7vtGK2KHo6uu7ga?dl=0

## Acknowledgements

The authors want to acknowledge help by Adriana Granich Armenta on data collection.

## Authors’ contributions

ACM conceived the research question. ACM, CMGL, and HLF designed the study and analysis plan. ACM and CMGL prepared the data and the statistical analysis. ACM, CMGL, MA, ACS drafted the initial and final versions of the manuscript. All authors critically reviewed early and final versions of the manuscript.

## Conflict of interest statements

The authors declared no conflict of interest.

## Role of funding source

This research received no funding. ACM was in grant period of COMEXUS-Fulbright while wrote this paper and received personal fees from the National Institute of Public Health of Mexico outside of the submitted work. Grants or personal fees had no role in study design, data collection and analysis, decision to publish, or preparation of the manuscript. The views expressed in this publication are those of the author(s) and not necessarily those of the institutions affiliated with.

**Supplementary Map 1.**
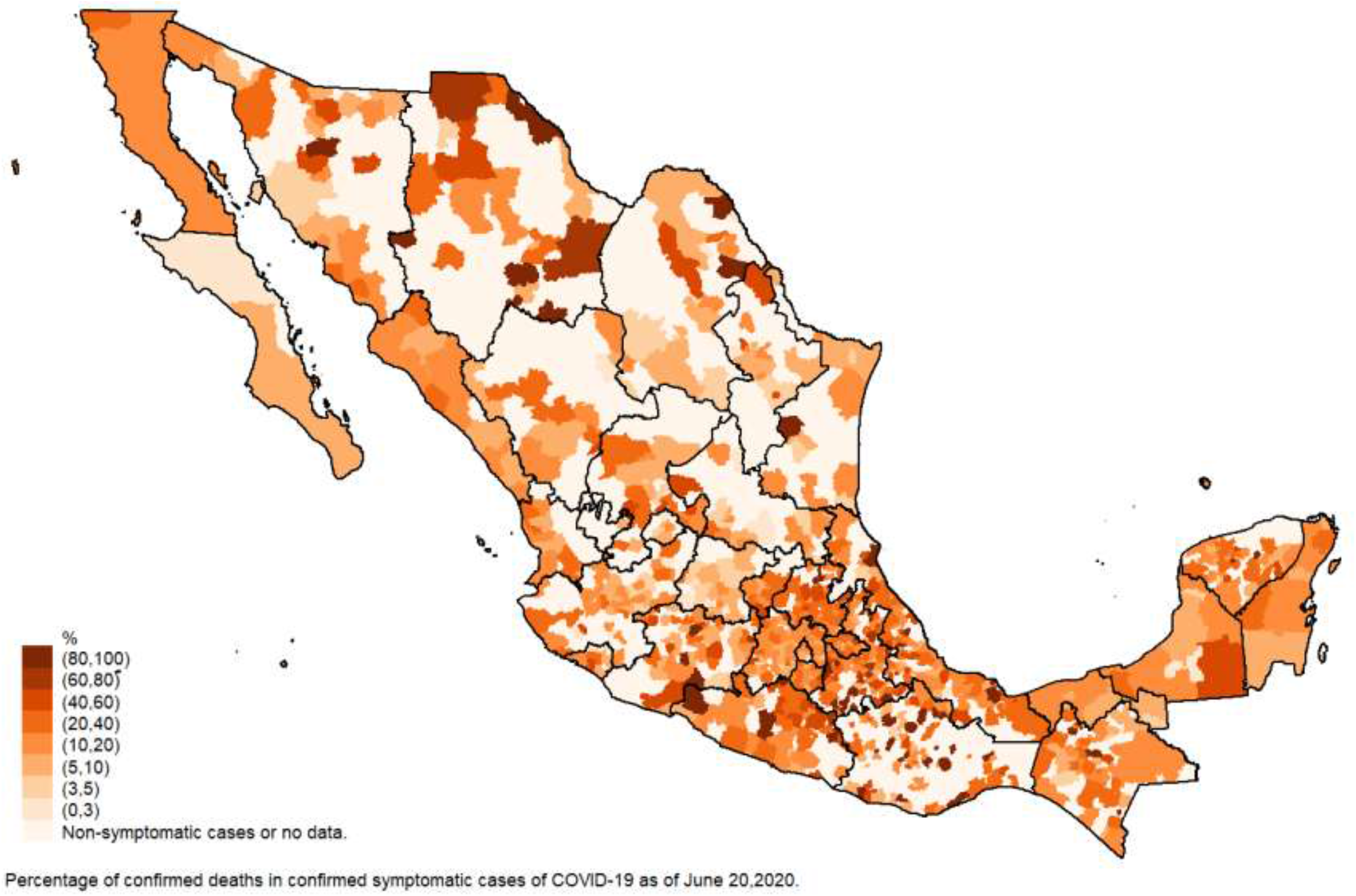
Case-fatality rate of COVID-19 in symptomatic cases.

## Notes

### Competing Interest Statement

The authors have declared no competing interest.

### Author Declarations

Our study did not involve human subjects.

